# Defining Core Competencies and Training Priorities for Infectious Disease Dynamics as a Discipline

**DOI:** 10.64898/2026.08.17.26360616

**Authors:** Lindsay T. Keegan, Kimberley Shoaf

## Abstract

Infectious disease dynamics is a growing, interdisciplinary field that aims to advance the understanding of how infectious diseases spread and how to control them. Most trainees enter the field through established disciplines and assemble *ad hoc* training and experience in infectious disease dynamics. As such, expectations for doctoral training remain largely implicit and highly variable across institutions. Other fields have formalized training expectations though defined training competencies, which promote transparency and alignment across institutions without prescribing specific approaches to training or research. In this paper, we set out to define the core competencies that characterize doctoral-level expertise in infectious disease dynamics. We assembled a team of seven people at the University of Utah and drafted a competency set. We then validated the competency set with experts in the field using an e-Delphi process. We did not restrict participation by location, job title, or sector. We set an a priori threshold for consensus to 70% and sent out two rounds of surveys to experts, asking them to rank the competencies by order of importance. Our team initially generated a list of 13 proposed Cross-cutting, 24 Applied Modeling, 17 Data Science, and 16 Theory competencies. After completing two rounds of validation, we validated two tracks comprised of 7 Cross-cutting, 10 Applied Modeling, and 12 Theory competencies. This study represents the first structured effort to define doctoral-level competencies in infectious disease that can help guide curriculum development, comprehensive exam preparation, and trainee evaluation, while also supporting alignment between academic training and workforce needs.

**Graphical abstract:** 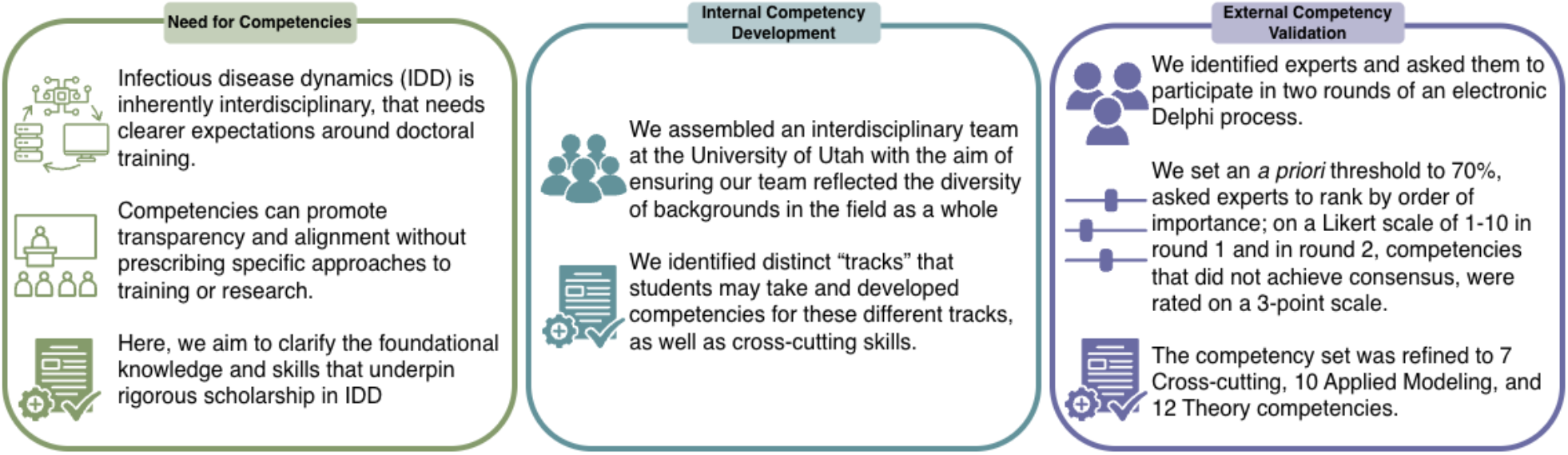

## Introduction

Over the past two decades, infectious disease dynamics (IDD) has emerged as a distinct interdisciplinary field that integrates mathematical and statistical theory, high-resolution data, and public health policy to advance the understanding of how infectious diseases spread and how to control them. While rooted in mathematical biology, the field has expanded alongside advances in computational power, genomic capabilities, and data collection resulting in the increasing reliance on models during major public health emergencies^1–4^ The COVID-19 pandemic accelerated this trajectory, driving rapid growth in research activity, training programs, and demand for scientists capable of working across theory, data, and applied public health practice.^5–9^ The establishment of the Global Society for Infectious Disease Dynamics (GSIDD) in January 2026 reflects the continued maturation of the field as a distinct scientific community.^10^

IDD is inherently interdisciplinary. The questions that define the field often require integration across epidemiology, ecology, mathematics, statistics, computation, biology, or the social context in which diseases spread.^1,4^ These problems span scales from within-host processes to population-level transmission and bridge theory and application, requiring researchers to synthesize perspectives across disciplines rather than train within a single parent field.^11^ Despite this maturation, formal training pathways in IDD have not kept pace with the field’s evolution. Most trainees enter through established disciplines such as epidemiology, mathematics, biostatistics, or biology and assemble IDD expertise through a combination of cross-disciplinary coursework, workshops, collaborations, and mentorship. As such, training experiences are shaped heavily by advisor expertise, departmental structure, and institutional resources, resulting in substantial variability in preparation across programs. As the field’s scope and influence have expanded the consequences of this ad hoc training structure have become more apparent. This variability highlights the need for clearly defined competencies to improve transparency, consistency, and ensure rigor in training across institutions.

In contrast to the mostly implicit expectations for doctoral training in IDD, other fields have formalized training expectations as they expanded and matured. In public health, accreditation standards require programs to demonstrate that graduates achieve defined competencies.^12,13^ Engineering programs accredited through ABET similarly articulate learning outcomes that define expectations in technical rigor, problem solving, and communication.^14^ Similarly, Professional organizations such as the Council of State and Territorial Epidemiologists (CSTE) have developed tiered competency frameworks to guide workforce development and clarify career expectations within public health practice.^15^ Together, these examples illustrate how clearly articulated competencies can promote transparency and alignment across institutions without prescribing specific approaches to training or research.

Articulating a competency framework would make explicit what has thus far remained implicit, clarifying foundational expectations for training and providing a shared structure for preparing trainees for careers spanning the full portfolio of IDD. Here, we set out to define the core competencies that characterize doctoral-level expertise in IDD. Drawing on structured expert input, we developed a consensus-based framework across cross-cutting, applied modeling, and theoretical domains. This effort is intended to clarify the foundational knowledge and skills that underpin rigorous scholarship in the field, rather than to standardize intellectual approaches or constrain disciplinary diversity.^16^ By articulating these competencies, we provide a foundation for strengthening training, fostering consistency across programs, and supporting the continued maturation of IDD as a discipline.

## Methods

To develop a competency set for IDD, we assembled a team of seven people at the University of Utah (Figure 1). This work was completed as part of our CDC-funded Healthcare, Infectious Diseases, Research (HIRe) Modeling Fellowship grant.^17^ The team included diverse perspectives from mathematics, epidemiology, biology, public health, internal medicine, computer science, and biostatistics, with the aim of ensuring our team reflected the diversity of backgrounds in the field as a whole. We met monthly for two years. Through these discussions, we determined that within the field there were distinct “tracks” that students may take and that these different tracks required different level of mastery for similar skills. Our team drafted and agreed upon cross cutting and track specific competencies.

**Figure 1.**
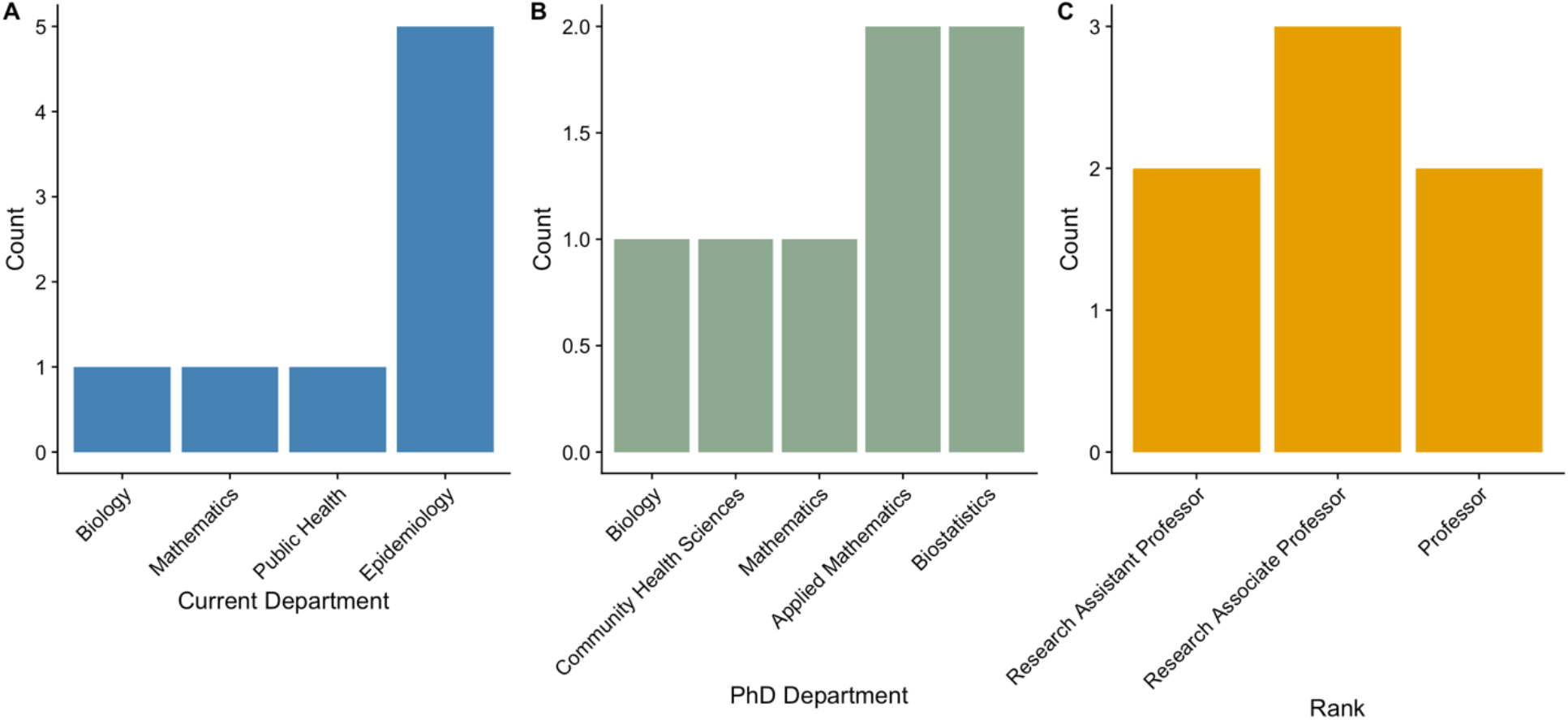
Demographics of our team at the University of Utah who developed the original competencies set (*see supplemental results*) by A) current department, B) department in which they got their PhD, and C) rank at the time of competency development.

We used a modified Delphi process^18–20^ to validate the competency set with experts in the field. While Delphi processes are generally conducted in person, ours was modified to be entirely via email and REDCap (also known as an e-Delphi or online Delphi).^18,21^ Experts were identified by members of our team, through other grantees of CDC awards including the HIRe fellowship program^17^ and its related program, the Modeling Infectious Diseases in Healthcare Network (MInD – Healthcare),^22^ through the CDC Center for Forecasting and Outbreak Analytics InsightNet,^23^ and through the Models of Infectious Disease Agent Study (MIDAS) network.^24^ We also used the snowball method to allow experts identified by experts we identified to participate.^25^ We did not restrict participation by location or job title and allowed US and international experts as well as academic and non-academic experts to respond. Figure 2 shows the demographic characteristics of respondents.

**Figure 2.**
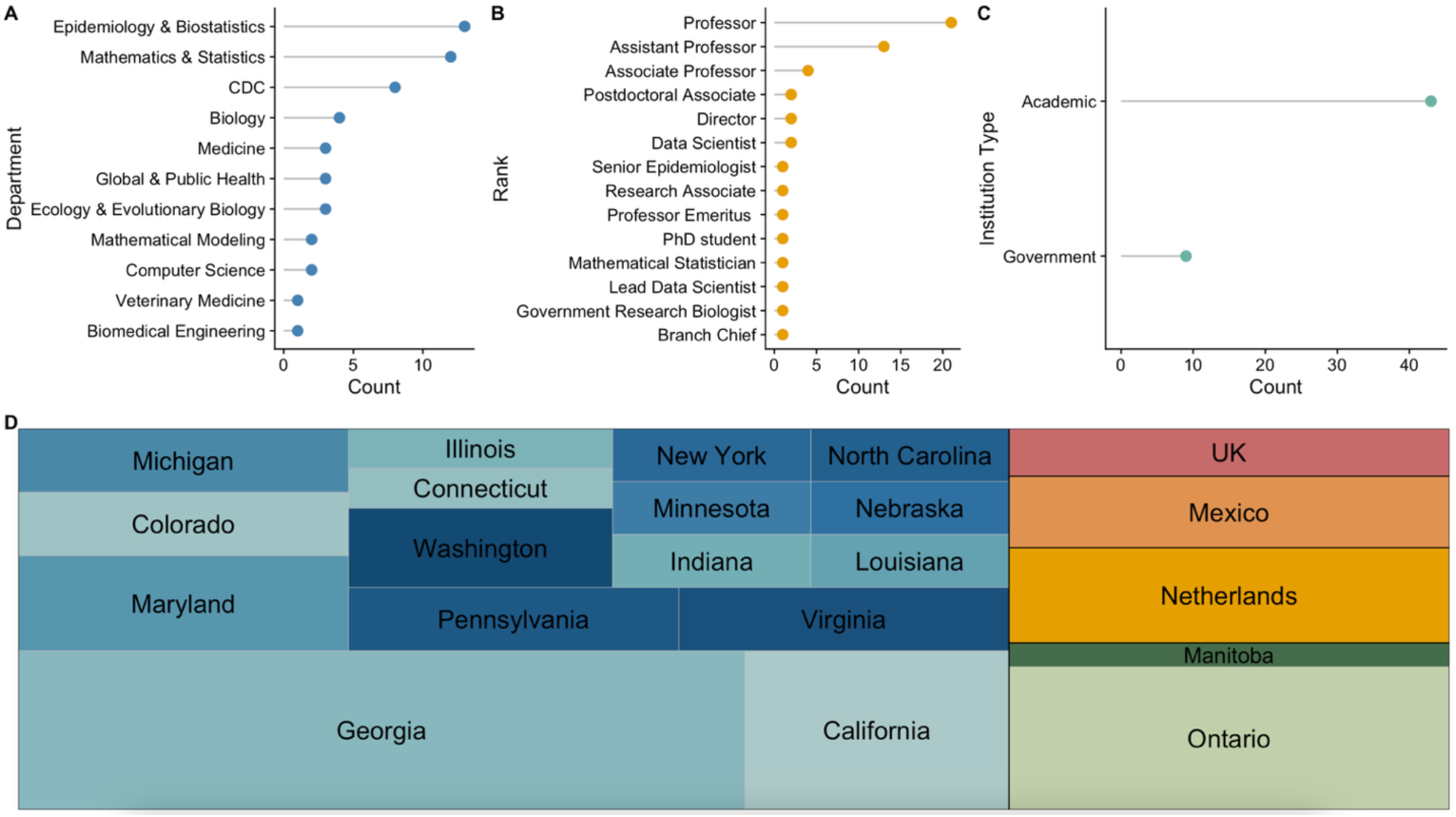
Demographics of respondents to the modified Delphi process who ranked the competencies set and determined the final set(*see supplemental results*) by A) department, B) rank at the time of responding, and C) institution type, and D) location, by state or province for the US and Canada or by country for the rest of the world.

We set an *a priori* threshold for consensus to 70% and sent out two rounds of surveys to experts, asking them to rank the competencies by order of importance. We asked all responders to evaluate cross-cutting competencies. Prior to answering track-specific questions, we asked experts to self-select their expertise and only asked them to rank track-specific questions for their self-identified expertise. In Round 1, respondents rated each item on a Likert scale of 1-10. Data were analyzed in Excel^26^ and R^27^, identifying items that met the consensus threshold. Competencies that did not achieve consensus were included in the second round of surveys, asking respondents to rate each item on a 3-point scale of 1) *Do Not Include*; 2) *Maybe Include*; 3) *Definitely Include*. Again, data were analyzed in Excel^26^ and R^27^, identifying items that met the consensus threshold.

## Results

### Initial Competency Generation

The TRANSMIT team initially generated a comprehensive list of proposed competencies across three domains including 13 Cross-cutting competencies, 24 Applied Modeling competencies, and 16 Theory competencies. These competencies were developed to reflect foundational knowledge, methodological rigor, and applied translational skills expected of doctoral-level trainees in IDD.

### Expert Participation

A total of 43 experts participated in the first round of the competency validation survey. Respondents self-identified their primary area of expertise as Applied Modeling (n = 28), Theory (n = 11), or Data Science (n = 4). A very limited number of responses were obtained from individuals with primary expertise in Data Science, therefore, we conducted targeted outreach following the first round to increase representation from this group; however, only four total responses were obtained in this domain. Due to the low response rate in Data Science, competencies in this category were excluded from the second round of validation and experts could only self-identify expertise in Applied Modeling or Theory; resulting in only two tracks for the final competency set. The second-round survey included 22 experts, with 15 identifying primarily with Applied Modeling and 7 with Theory.

### Competency Validation and Consensus

Following two rounds of expert review and validation, the competency set was refined to 7 Cross-cutting competencies, 10 Applied Modeling competencies, and 12 Theory competencies. Reductions reflected consolidation of overlapping items, clarification of scope, and removal of competencies that did not achieve consensus regarding their centrality to doctoral-level training in IDD. A full list of the pre-validation and validated competency sets can be found in the supplemental results.

### Examples of validated and cut competencies

To give some examples of the competencies that were validated and cut by experts, we selected representative competencies from each of the tracks. For cross-cutting competencies, experts agreed that that doctoral trainees should be able to “*read and critically evaluate relevant scientific literature in infectious disease dynamics*” and “*describe the application of models of your work/science to policy or practice*”; from the applied modeling track, experts agreed that that doctoral trainees should be able to “*describe the applications and limitations of infectious disease predictive modeling in preparedness planning, forecasting, and policy guidance*,” “*demonstrate the ability to evaluate your assumptions (e*.*g*., *whether your work is realistic)*”, and “*define the model and evaluate whether it is using an ideal situation or real-world situations*;” and finally from the theory track, experts agreed doctoral students should be able to “*formulate and solve at least one type of mathematical problem derived from an epidemiological model (e*.*g*., *solving a differential equation, identifying a bifurcation point, or calculating a reproduction number)*” and “*produce a work that advances the theory of modeling in your area of expertise/field*.” These competencies reflect shared expectations regarding analytic rigor, theoretical grounding, and translational awareness.

In contrast, several proposed competencies either did not achieve consensus or achieved consensus to exclude them from the final framework. An example of one of the competencies that were cut from each of the tracks are, from the cross-cutting competencies: “*demonstrate the ability to navigate the publication process, including selecting an appropriate journal and determining when a manuscript is ready for submission*”; from the applied modeling track: “*identify populations experiencing health inequities and describe at least two approaches for addressing inequities within modeling frameworks*”; from the theory track: “*demonstrate proficiency in dynamic reporting tools (e*.*g*., *Quarto, R Markdown)*.” In general, rejected competencies were viewed as either too specialized, overly context-dependent, or insufficiently central to defining the core intellectual and methodological identity of IDD at the doctoral level.

## Discussion

This study represents the first structured effort to define doctoral-level competencies in IDD. Through iterative expert review, we refined an initial set of competencies to a consensus-driven framework across three domains: Cross-cutting, Applied modeling, and Theory. The consensus emphasizes shared expectations around analytic rigor, theoretical grounding, and translational awareness over technical tools or professional mechanics. This competency framework provides shared expectations without prescribing a single curriculum and clarifies expectations for what defines fluency in IDD at the doctoral level.

We believe the final competency set reflects the intellectual identity of IDD, with a strong emphasis on conceptual and mathematical rigor, critical evaluation, and translational relevance. Across domains, the framework prioritizes the ability to formulate, construct, and solve models derived from real-world epidemiological systems while rigorously evaluating assumptions, model quality, and appropriateness for available data. The competencies highlight the importance of integrating theory and application, requiring trainees to engage in both theoretical model development and simulation-based, programming-driven analysis. Equally central is the capacity to compare and interpret model outputs in light of their assumptions and to articulate the applications and limitations of modeling for preparedness planning and policy guidance. The framework also underscores the need for effective scientific communication across audiences and disciplines, foundational understanding of infectious disease biology relevant to modeling work, and the expectation that doctoral trainees contribute original scholarship that advances theory, methodology, or applied modeling within their area of expertise. The final competency set emphasizes rigor, integration across domains, and an explicit understanding of how models inform practice.

The competencies that did not achieve consensus are also informative. Many of the excluded items focused on professional mechanics including navigating the publication process, project management, or organizing deliverables. These skills matter for academic success, but they are not unique to IDD. Other rejected items were tied to specific software platforms or reporting tools. Reviewers appeared to favor durable analytic principles over proficiency in particular technologies that will inevitably change.

Several proposed competencies related to health equity were also not retained. More prescriptive formulations, such as requiring identification of specific populations experiencing inequities or enumerating particular approaches to incorporating equity into models, did not achieve consensus. While we believe that this does not reflect a lack of importance of equity in IDD, it may reflect a preference for framing these issues through general modeling principles rather than through specified procedural requirements. Fundamentally, health equity is a question of heterogeneity: variation in risk, contact patterns, access to care, and exposure. Indeed, while competencies related to health equity specifically did not achieve consensus, the more general concept of heterogeneity is embedded throughout the retained competencies, particularly those related to model construction, evaluation of assumptions, and interpretation in real-world settings. The reviewers appeared to favor integrating equity through this broader treatment of heterogeneity within core modeling practice rather than specifying detailed equity requirements at the competency level. Models are built on variation, and that expectation is reflected in the competency framework.

Several limitations should be considered when interpreting this framework. First, overall response counts were modest, and representation across subdomains was uneven, with particularly low participation from experts identifying primarily with data science. Despite targeted outreach following the first survey round, response rates in this area remained limited, potentially constraining the breadth of perspectives reflected in the final competency set. Second, the respondent pool was composed largely of academics, which may introduce disciplinary or institutional bias in defining doctoral-level expectations. Third, the framework reflects current disciplinary priorities at a particular moment in the field’s development and will likely require revision as methodological advances, technologies, and societal needs evolve. Additionally, competencies were evaluated at a relatively high level of abstraction; specific assessment metrics or performance benchmarks were not defined and remain an important area for future work. Finally, the framework is not intended to capture every specialized subfield within IDD, but rather to articulate shared foundational expectations applicable across diverse research trajectories.

As other disciplines have matured, they have clarified expectations for training through explicitly articulated competencies; IDD has reached a similar point. The framework proposed here reflects an effort to define what rigorous skills and knowledge doctoral-level preparation in the field of IDD should encompass. It captures the interdisciplinary structure of our field and helps to distinguish it from its parent disciplines. Our intent is not to prescribe a uniform curriculum, but to make expectations of skills and knowledge explicit. In practice, we hope the framework will be used to shape curriculum, guide comprehensive exam expectations, and help programs assess whether core areas are adequately covered. It also offers trainees and advisors a shared foundation for evaluating preparation as the field continues to evolve.

This framework represents an initial step toward clarifying doctoral-level expectations in IDD and invites further refinement. Development of structured assessment tools aligned with the proposed competencies would help to facilitate practical implementation within doctoral programs. Periodic revision will be essential as the discipline continues to evolve in response to new methodological innovations and public health challenges. Additionally, we hope that this can serve as a reference for developing competencies for other career stages such as master’s-level training, postdoctoral expectations, and workforce pathways, allowing for clearer alignment across stages of professional development. Empirical evaluation of whether competency-based training improves preparedness, scholarly productivity, and career outcomes would further strengthen the evidence base for this approach.

## Data Availability

All data produced in the present work are contained in the manuscript

## Declaration of generative AI use

We did not use generative AI.

## Data statement

Data are available at: https://github.com/UT-IDDynamics

## Acknowledgements

We acknowledge Frederick Adler, Yue Zhang, Damon Toth, Karim Khader, and George Vega Yon for their help constructing the initial competency set and Hannah Higgs for her help with project management and REDCap administration.

## SUPPLEMENTAL INFORMATION

### Supplemental Results

#### Pre-validation competency set REDCap Survey

The following competency statements are considered core or cross-cutting competencies that are relevant for all 3 fields of modelers. Please read each statement and provide your rating of how important it is that modelers across all 3 fields have that competency. Rate each one on a scale of 0 (Not at all important) to 10 (extremely important).

1. Demonstrate what it means to be a good mentor/mentee.
2. Construct a body of work that advances your area of expertise.
3. Read and understand relevant literature.
4. Communicate complex information clearly in figures that are accessible to all in the scientific community (e.g., color choice).
5. Demonstrate effective written communication and presentation skills within your field.
6. Communicate across disciplines and across settings (e.g., academia, public health, corporate, students).
7. Demonstrate the ability to navigate the publication process including choosing an appropriate journal and determining when a paper is done and ready to submit.
8. Understand the fundamentals of project management.
9. Demonstrate ability to break down work into tasks and create a feasible schedule.
10. Describe a project’s constraints in scope, schedule, budget, risk, quality, and resources.
11. Describe and track project deliverables.
12. Describe the application of models of your work/science to policy or practice (broader impacts).
13. Describe the principles of health equity.
14. Discuss how to include equity in models, or challenges for specific areas/populations.
15. Explain disease systems / biology of disease relevant to the student’s area of focus (e.g., biology of modeling evolution, ecology, microbiology, immunology, healthcare settings and treatment/interventions).

### If you work in the theory field, please respond to the following

1. Describe multiple types of mathematical models used in infectious disease epidemiology (e.g., compartmental, agent-based, stochastic).
2. Formulate solution for at least one type of mathematical problem derived from an epidemiological model (e.g., solve differential equation, find bifurcation point, calculate reproduction number).
3. Evaluate model for appropriateness for use of real-world data.
4. Construct a theoretical mathematical model of one real-world epidemiological system.
5. Produce a work that advances the theory of modeling in your area of expertise/field.
6. Demonstrate the ability to evaluate your assumptions (e.g., whether your work is realistic). Define the model and evaluate whether it is using an ideal situation or real-world situations.
7. Write a review manuscript.
8. Describe the applications and limitations of infectious disease predictive modeling in preparedness planning, forecasting, and guidance for policy makers.
9. Compare and interpret the results of different infectious disease models and scenarios, taking into account their assumptions.
10. Demonstrate a basic understanding of advanced technologies for reproducibility, namely, version control software (Git+GitHub) and Containers (Docker).
11. Demonstrate the ability to use dynamic reporting: quarto.org and RMarkdown.
12. Demonstrate the fundamentals of data visualization using ggplot2 and plotly (https://plotly.com/r/.) The latter is for interactive data visualization. This is connected to the previous point. It should also discuss color blindness.
13. Demonstrate the ability to perform simulation-based modeling
14. Demonstrate awareness of MATLab, R, Students should be aware of available software/programming language (e.g., MATLab, R)
15. Use one or more techniques for programming infectious disease models.
16. Demonstrate the use of reticulate python.

### If you work in the data science field, please respond to the following

1. Employ multiple types of models (e.g., compartmental, agent-based, stochastic, time series, network) and describe their uses (e.g., parameter estimation, forecasting, nowcasting).
2. Assess model quality.
3. Evaluate your assumptions, by defining the model and evaluating whether it is using an ideal real-world situation.
4. Write a review manuscript.
5. Describe the applications and limitations of infectious disease predictive modeling in preparedness planning, forecasting, and guidance for policy makers.
6. Compare and interpret the results of different infectious disease models and scenarios, taking into account their assumptions.
7. Demonstrate the ability to perform simulation-based modeling.
8. Demonstrate a basic understanding of advanced technologies for reproducibility, namely, version control software (Git+GitHub) and Containers (Docker).
9. Demonstrate the ability to use dynamic reporting: quarto.org and RMarkdown.
10. Demonstrate the fundamentals of data visualization using ggplot2 and plotly (https://plotly.com/r/.) The latter is for interactive data visualization.
11. Use one or more techniques for programming infectious disease models.
12. Demonstrate awareness of MATLab, R, Students should be aware of available software/programming language (e.g., MATLab, R).
13. Demonstrate the use of reticulate python.
14. Conduct an analysis of models.
15. Conduct data wrangling using basic statistics and maximum likelihood and Bayesian methods to link data with dynamical models.
16. Demonstrate a basic understanding of machine learning and deep learning.
17. Identify a variety of data streams - e.g., novel publicly available data streams.

### If you work in the applied modeling field, please respond to the following

1. Evaluate multiple types of models (e.g., compartmental, agent-based, stochastic) and their uses (e.g., parameter estimation, forecasting, nowcasting)
2. Assess model quality.
3. Evaluate your assumptions, by defining the model and evaluating whether it is using an ideal real-world situation.
4. Write a review manuscript.
5. Describe the applications and limitations of infectious disease predictive modeling in preparedness planning, forecasting, and guidance for policy makers.
6. Compare and interpret the results of different infectious disease models and scenarios, taking into account their assumptions.
7. Demonstrate the ability to perform simulation-based modeling.
8. Demonstrate a basic understanding of advanced technologies for reproducibility, namely, version control software (Git+GitHub) and Containers (Docker).
9. Demonstrate the ability to use dynamic reporting: quarto.org and RMarkdown.
10. Demonstrate the fundamentals of data visualization using ggplot2 and plotly (https://plotly.com/r/.) The latter is for interactive data visualization. This is connected to the previous point. It should also discuss color blindness.
11. Demonstrate awareness of MATLab, R, Students should be aware of available software/programming language (e.g., MATLab, R).
12. Use one or more techniques for programming infectious disease models.
13. Demonstrate the use of reticulate python.
14. Conduct analysis of models.
15. Translate the results of your work into a format for policy makers (e.g., white paper).
16. Enumerate the different levels of governmental public health
17. Describe the decision making authority of the players involved in making public health decisions
18. Describe at least 2 potential groups of people impacted by those decisions.
19. Enumerate the non-governmental entities who might use results of models for decisions (e.g., hospitals, health systems, etc).
20. Identify potential populations who experience health inequities and describe at least two approaches to handling those inequities in models.
21. Communicate about models and results to at least two populations (community, public health professionals, elected officials, etc.).
22. Describe the application of models of your work/science to policy or practice (broader impacts).
23. List the different stakeholders where decisions can be made, and who those decisions could impact.
24. Discuss how to include equity in models, or challenges for specific areas/populations.

**From your perspective are there any competencies missing from the list?** Please write any competencies that you think should be added. Please note whether the added competency would be considered a core/cross-cutting competency or for one of the fields (please note the field).

#### Validated competency set

##### Core/Cross-Cutting

1. Construct a body of work that advances your area of expertise.
2. Read and understand relevant literature.
3. Communicate complex information clearly in figures that are accessible to all in the scientific community (e.g., color choice).
4. Demonstrate effective written communication and presentation skills within your field.
5. Communicate across disciplines and across settings (e.g., academia, public health, corporate, students).
6. Describe the application of models of your work/science to policy or practice (broader impacts).
7. Explain disease systems / biology of disease relevant to the student’s area of focus (e.g., biology of modeling evolution, ecology, microbiology, immunology, healthcare settings and treatment/interventions).

##### Applied Modeling

1. Evaluate multiple types of models and their uses (e.g., parameter estimation, forecasting, nowcasting)
2. Assess model quality.
3. Evaluate your assumptions, by defining the model and evaluating whether it is using an ideal real-world situation.
4. Describe the applications and limitations of infectious disease predictive modeling in preparedness planning, forecasting, and guidance for policy makers.
5. Compare and interpret the results of different infectious disease models and scenarios, taking into account their assumptions.
6. Demonstrate the ability to perform simulation-based modeling.
7. Use one or more techniques for programming infectious disease models.
8. Conduct analysis of models.
9. Describe the application of models of your work/science to policy or practice (broader impacts).
10. Discuss how to include equity in models, or challenges for specific areas/populations.

##### Theory

1. Describe multiple types of mathematical models used in infectious disease epidemiology (e.g., compartmental, agent-based, stochastic).
2. Formulate solution for at least one type of mathematical problem derived from an epidemiological model (e.g., solve differential equation, find bifurcation point, calculate reproduction number).
3. Evaluate model for appropriateness for use of real-world data.
4. Construct a theoretical mathematical model of one real-world epidemiological system.
5. Produce a work that advances the theory of modeling in your area of expertise/field.
6. Demonstrate the ability to evaluate your assumptions (e.g., whether your work is realistic).
7. Define the model and evaluate whether it is using an ideal situation or real-world situations.
8. Describe the applications and limitations of infectious disease predictive modeling in preparedness planning, forecasting, and guidance for policy makers.
9. Compare and interpret the results of different infectious disease models and scenarios, taking into account their assumptions.
10. Demonstrate the ability to perform simulation-based modeling
11. Demonstrate awareness of MATLab, R, Students should be aware of available software/programming language (e.g., MATLab, R)
12. Use one or more techniques for programming infectious disease models.

